# DNA methylation outlier burden, health and ageing in Generation Scotland and the Lothian Birth Cohorts of 1921 and 1936

**DOI:** 10.1101/19010728

**Authors:** Anne Seeboth, Daniel L. McCartney, Yunzhang Wang, Robert F. Hillary, Anna J. Stevenson, Rosie M. Walker, Kathryn L. Evans, Andrew M. McIntosh, Sara Hägg, Ian J. Deary, Riccardo E. Marioni

## Abstract

DNA methylation outlier burden has been suggested as a potential marker of biological age. An outlier is typically defined as DNA methylation levels at any one CpG site that are three times beyond the inter-quartile range from the 25^th^ or 75^th^ percentiles compared to the rest of the population. DNA methylation outlier burden (the number of such outlier sites per individual) increases exponentially with age. However, these findings have been observed in small samples. Here, we showed an association between age and log10-transformed DNA methylation outlier burden in a large cross-sectional cohort, the Generation Scotland Family Health Study (N=7,010, β=0.0091, p<2×10^−16^), and in two longitudinal cohort studies, the Lothian Birth Cohorts of 1921 (N=430, β=0.033, p=7.9×10^−4^) and 1936 (N=898, β=7.9×10^−3^, p=0.074). Significant confounders of both cross-sectional and longitudinal associations between outlier burden and age included white blood cell proportions, BMI, smoking, and batch effects. In Generation Scotland, the increase in epigenetic outlier burden with age was not purely an artefact of an increase in DNA methylation level variability with age (epigenetic drift). Log10-transformed DNA methylation outlier burden in Generation Scotland was not related to self-reported, or family history of, age-related diseases and it was not heritable (SNP-based heritability of 4.4%, p=0.18). Finally, DNA methylation outlier burden was not significantly related to survival in either of the Lothian Birth Cohorts individually but it was in a meta-analysis (HR_meta_=1.12; 95%CI_meta_=[1.02; 1.21]; p_meta_=0.021). These findings suggest that, while it does not associate with ageing-related health outcomes, DNA methylation outlier burden does track chronological ageing and may also relate to survival. DNA methylation outlier burden may thus be useful as a marker of biological ageing.

## Background

Chronological age is a major risk factor for many diseases. Consequently, as life expectancy increases, so too does the prevalence of age-related diseases. However, there are considerable inter-individual differences in health outcomes (1). In order to improve risk stratification beyond crude models based on chronological age, much recent work has thus attempted to identify and test the utility of markers of ‘biological age’.

One focus of research into ageing biomarkers has been on age-related changes in DNA methylation at cytosine-guanine (CpG) dinucleotide sites. DNA methylation is a dynamic epigenetic modification that is involved in the regulation of gene expression, and is influenced by both genetics (2) and the environment (3). Analyses have identified a number of loci where methylation changes consistently across individuals as they age, for example the cg16867657 locus in the ELOVL2 gene (4). In addition to such sites, methylation levels at other loci have been shown to diverge as individuals grow older (5). This is consistent with the observation that, on average, inter-individual variability in DNA methylation tends to increase as people get older, a phenomenon referred to as epigenetic drift (5,6).

Recently, researchers have started to investigate the accumulation of DNA methylation outliers (sometimes referred to as ‘stochastic epigenetic mutations’). DNA methylation outliers occur when the DNA methylation level at a specific site in an individual’s genome differs greatly from that of the majority of the population at this locus. It is known that such unusual methylation levels can lead to aberrant gene expression, and DNA methylation outliers have been shown to be associated with the development of certain cancers (7,8) or neurodevelopmental disorders and congenital anomalies (9).

In addition, many studies have investigated the dysregulation of biological processes with age. This includes innate immune system function (10) but also epigenetic processes such as DNA methylation (11). DNA methylation outlier burden (i.e. the number of outlier sites per individual), as a novel measure of such dysregulation, may thus also have predictive value as an index of biological ageing.

Indeed, Gentilini and colleagues (12) found an exponential association between age and DNA methylation outlier burden in 170 individuals aged between 3 and 106 years (r_log (outlier burden) – age_ = 0.63). The authors defined outliers as methylation levels of greater than 3 times the inter-quartile range (IQR) from the 25^th^ and 75^th^ percentiles compared to the rest of the population. More recently, two further studies replicated this finding using data from 658 individuals of a wide age range (mean=54.3 years, SD=12.7 years) and longitudinal data from 375 older individuals (48 to 98 years), respectively (13,14). Both studies reported individual outlier burden to be highly variable (119-18,308 out of 769,042 CpGs (14); range 58-26,291 out of 370,234 CpGs (13)) and non-normally distributed. Following log-transformation, these studies found outlier burden to be higher in older individuals (z=8.54; p=1.2×10^−17^ (14); β=8.3×10^−3^, p=1.2×10^−13^ (13)). In addition, white blood cell proportions were found to be significantly associated with outlier burden in both studies.

Here, we provide a comprehensive characterisation of DNA methylation outlier burden in a large cross-sectional sample of 7,010 individuals and in two longitudinal samples of 430 and 898 individuals, respectively. We investigate the association of DNA methylation outlier burden with age, explore the potential confounding effect of epigenetic drift on these findings, relate outlier burden to more than a dozen health- and ageing-related traits and to survival, and we determine the genetic contribution to individual differences in outlier burden.

## Results

### Descriptive Statistics

Descriptive statistics for age, sex and outlier burden in the three cohorts are shown in Table 1 (for more detailed descriptive statistics including covariate information, see Supplementary Tables 1A, 1B and 1C, Additional File 1).

**Table 1.** Descriptive Statistics.

|  | **Wave 1** | **Wave 2** | **Wave 3** | **Wave 4** |
| --- | --- | --- | --- | --- |
| **GS:SFHS** |  |  |  |  |
| **N** | 7010 (100%) | - | - | - |
| **Females** | 4076 (58%) | - | - | - |
| **Age (years)** | 51 (13) | - | - | - |
| **Outlier Burden** | 1119 (3361) | - | - | - |
| **LBC1921** |  |  |  |  |
| **N** | 430 (100%) | - | 173 (100%) | 82 (100%) |
| **Females** | 260 (60%) | - | 94 (54%) | 44 (54%) |
| **Age (years)** | 79 (0.58) | - | 87 (0.40) | 90 (0.10) |
| **Outlier Burden** | 2039 (3280) | - | 2330 (3901) | 1717 (2029) |
| **LBC1936** |  |  |  |  |
| **N** | 898 (100%) | 793 (100%) | 607 (100%) | 502 (100%) |
| **Females** | 445 (50%) | 376 (47%) | 291 (48%) | 249 (50%) |
| **Age (years)** | 70 (0.83) | 73 (0.70) | 76 (0.67) | 79 (0.62) |
| **Outlier Burden** | 2197 (3203) | 2729 (6020) | 2293 (6861) | 1423 (2845) |

*Note.* Sample numbers (N) alongside their proportion in the entire sample (in %) are reported for categorical variables. Mean (M) and Standard Deviation (SD) values are reported for continuous variables.

In Generation Scotland, the mean number of outliers per individual was 1,119 (SD=3,361) and highly variable, ranging from 53 to 84,933 (out of 334,352 and 349,027 CpG sites in set 1 and set 2, respectively). The distribution of outlier number per individual was positively skewed, even after log10-transformation. In the combined Generation Scotland dataset 97% of 361,846 CpG sites had at least one individual with a DNA methylation outlier at this site. On average, CpG sites had 22 individuals with DNA methylation outliers at the site (SD=33, max=1,587, across 7,010 individuals). In the LBC1921 and in the LBC1936 at wave 1, there were on average 2,039 (min=108; max=27,676; SD=3,280) and 2,197 (min=119; max=35,711; SD=3,203) DNA methylation outliers per individual (out of 356,631 CpG sites), respectively. The distribution of outlier number per individual was positively skewed. Following log10-transformation, the distribution looked approximately normal. Sixty-three percent of 356,631 CpG sites were found to have at least one individual with an outlier at this site in the LBC1921 at wave 1 (83% in the LBC1936). On average, CpG sites had three individuals with outliers a this site (SD=5, max=108, across 430 individuals) in the LBC1921 and six individuals with outliers at this site (SD=10, max=225, across 898 individuals) in the LBC1936.

### DNA methylation outlier burden and age

In Generation Scotland, there was a small but significant cross-sectional association between age and log10(outlier burden) in all models regardless of adjustments (Supplementary Table 2, Additional File 1) (β_raw_=9.1×10^−3^, p<2×10^−16^, fully adjusted model; this corresponded to a 2.1% higher outlier burden per year older in age). The association between log10(outlier burden) and age was non-linear (Figure 1). There was evidence of heteroscedasticity in the model residuals (Supplementary Figure 1, Additional File 1).

**Figure 1.**
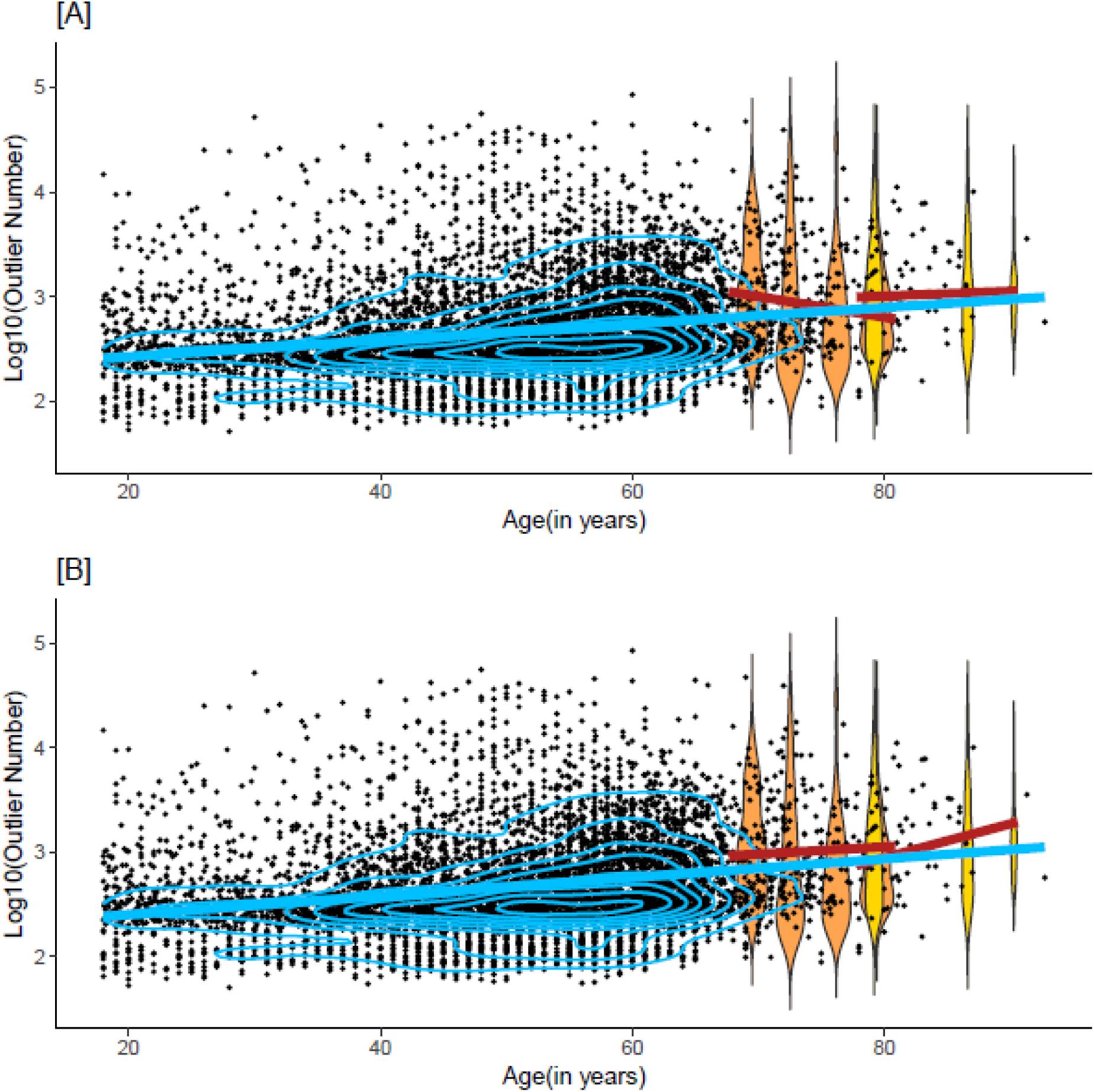
DNA methylation outlier burden in Generation Scotland, LBC1921 and LBC1936. Distribution of log10 (outlier burden) in Generation Scotland (black, blue contour shapes indicating data density) and in the four LBC1936 waves (orange) and three LBC1921 waves (yellow). Panel A shows linear regression lines in Generation Scotland (blue) and in the LBC1921 and LBC1936 (red) to model the association of *log10(Outlier Burden) ∼ age*. Panel B shows fitted values for the regression of *log10(outlier burden) ∼ age* with random factor batch and cell proportions fit to the mean.

A number of covariates included in the fully adjusted model were significantly associated with DNA methylation outlier burden (Supplementary Table 3A, Additional File 1). B-cell, NK, CD8^+^ T-cell and granulocyte proportions were positively associated with outlier burden (β_raw_=0.15, p<2×10^−16^; β_raw_=0.075, p=6.6×10^−10^; β_raw_=0.084, p=2.2×10^−16^ and β_raw_=0.045, p=0.026; coefficients refer to a unit change in log10(outlier burden) per standard deviation change in the predictor). The proportion of CD4^+^ T-cells was negatively associated with outlier burden (β_raw_=-0.078, p=4.5×10^−9^). Log10(pack years +1) was positively associated and log10(BMI) was negatively associated with outlier burden (β_raw_=0.025, p=4.3×10^−3^; β_raw_=-0.013, p=8.0×10^−3^). Sex, never vs. ever smoking status and cancer were not significantly associated with outlier burden.

Longitudinally, and prior to adjustments, DNA methylation outlier burden increased with age in LBC1921 and decreased with age in the LBC1936 (Figure 1; Figure 2). However, after adjustment for covariates, the association between age and log10(outlier burden) was positive in both cohorts (Figure 1). This association was significant after adjustment for cell counts in the LBC1921 (β_raw_=0.033, p= p=7.9×10^−4^, fully adjusted model; this corresponded to a 7.9% increase in outlier burden per year increase in age) but not in the LBC1936 (β_raw_=7.9×10^−*3*^, p=0.074, fully adjusted model; 1.8% increase per year) (Supplementary Table 2, Additional File 1). Ageing trajectories for those individuals with complete observations, i.e. observations in all waves (63 in the LBC1921 and 337 in the LBC1936), were similar to those observed in the entire sample (Supplementary Figures 2, 3 and 4, Additional File 1).

**Figure 2.**
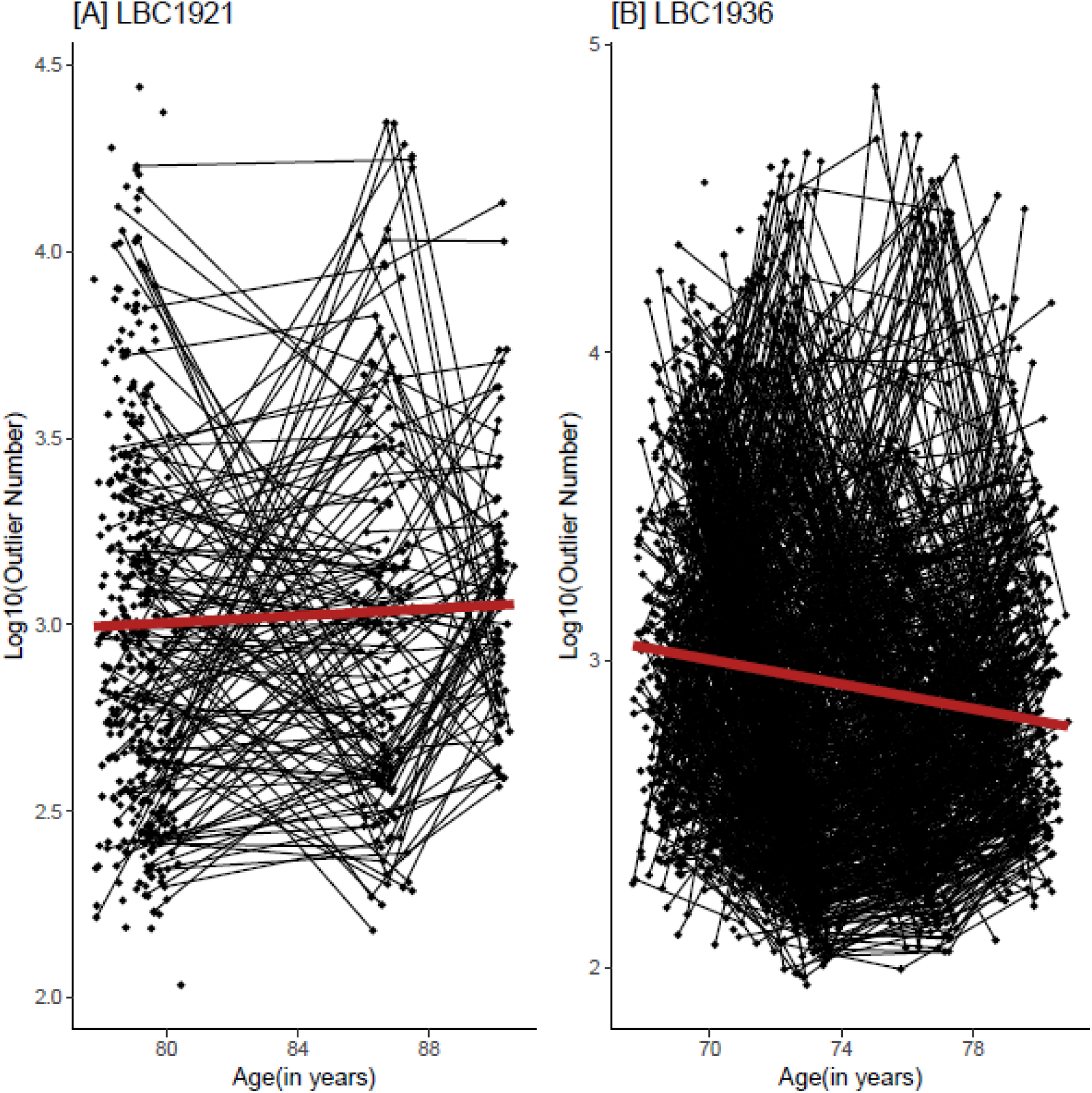
Trajectories of DNA methylation outlier burden in the LBC1921 and LBC1936. Longitudinal change in log10(outlier burden) in individuals in the LBC1921 and the LBC1936. The linear regression lines of *log10(Outlier Burden) ∼ age* are shown in red.

As in Generation Scotland, estimated cell proportions of B-cells, NK cells and CD8^+^ T-cells in the LBC1921 (β_raw_=0.20, p<2×10^−16^; β_raw_=0.13, p=6.7×10^−6^; β_raw_=0.082, p=2.8×10^−3^) and B-cells, NK cells and CD4^+^T-cells in the LBC1936 were significantly related to outlier burden (β_raw_=0.13, p<2×10^−16^; β_raw_=0.056, p=1.0×10^−4^; β_raw_=-0.059, p=1.1×10^−3^). No other covariates were significantly associated with DNA methylation outlier burden in either the LBC1921 or the LBC1936 (Supplementary Table 3B and 3C, Additional File 1).

### DNA methylation outlier burden and epigenetic drift

In Generation Scotland, DNA methylation outlier burden based on an alternative definition of outliers within age groups correlated almost perfectly with outlier burden calculated using the original definition (r=0.99, p<2×10^−16^). Accounting for epigenetic drift by applying the alternative definition for outliers slightly attenuated the association between DNA methylation outlier burden and age in Generation Scotland (Supplementary Table 4A, Additional File 1) (β_raw_=6.2×10^−3^, p<10^−16^, fully adjusted model; corresponding to a 1.4% higher DNA methylation outlier burden per year older in age). However, the association was still highly significant and the pattern of covariate associations did not change (effect sizes for each covariate presented in Supplementary Table 4B, Additional File 1).

### DNA methylation outlier burden, health and survival

We did not find evidence for a cross-sectional association between DNA methylation outlier burden and self-reported disease (Supplementary Tables 5A and 5B, Additional File 1) or family history of disease (Supplementary Tables 6A and 6B, Additional File 1) in Generation Scotland following Bonferroni correction.

DNA methylation outlier burden was not significantly associated with survival in the LBC1921 or the LBC1936 individually (Figure 3; Supplementary Table 7, Additional File 1). However, when meta-analysed across the two cohorts, the association between outlier burden and survival was significant at a nominal P<0.05 threshold (HR_meta_=1.12; 95%CI_meta_=[1.02; 1.21]; p_meta_=0.021).

**Figure 3.**
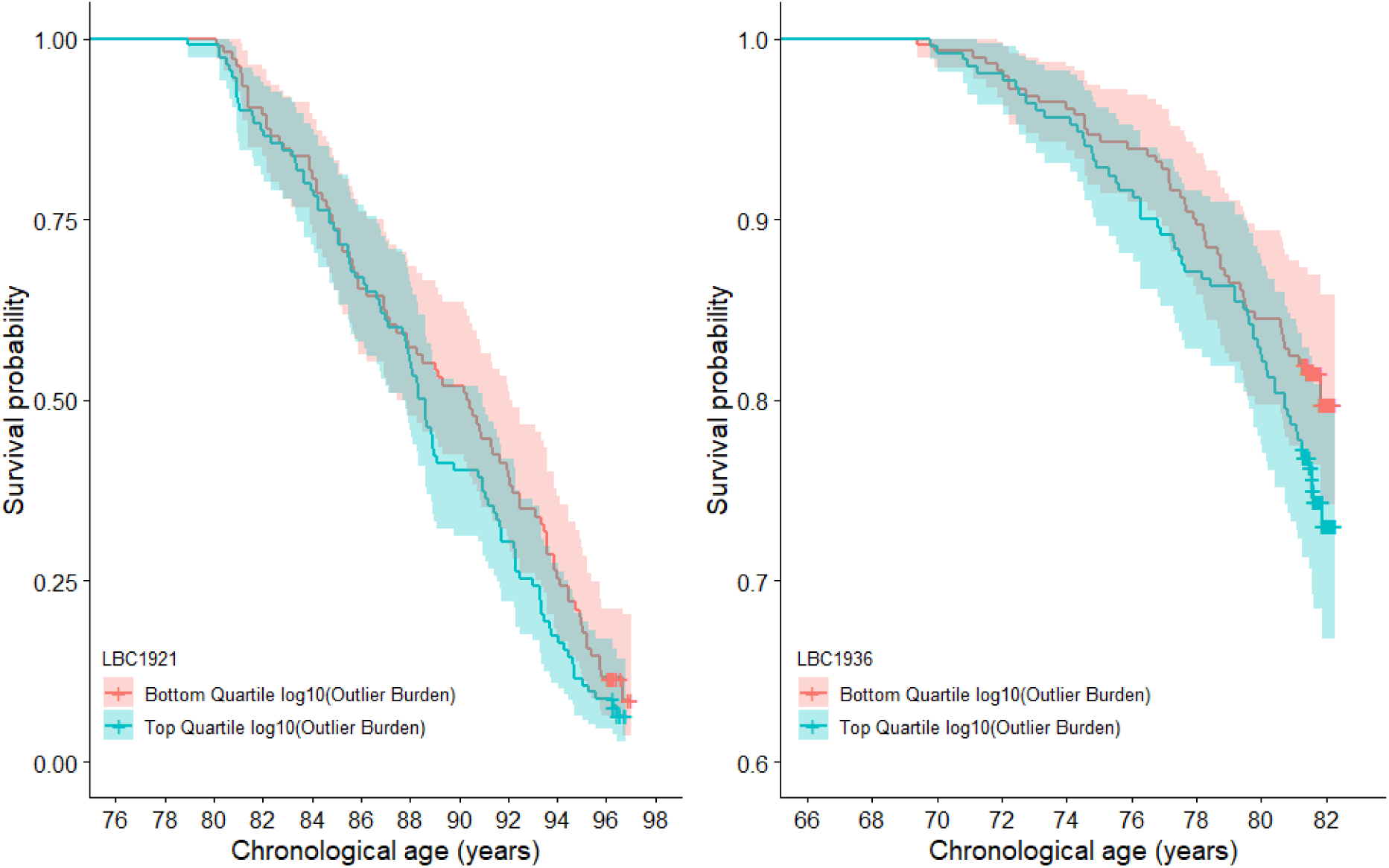
DNA methylation outlier burden survival plot in the LBC1921 and the LBC1936. Survival probability by top and bottom quartile of log10(outlier burden) adjusted for sex and chronological age in the LBC1921 and in the LBC1936.

### Heritability of DNA methylation outlier burden

There was no significant genetic contribution by all common genome-wide SNPs (minor allele frequency > 5%) to phenotypic variability in log10—transformed DNA methylation outlier burden (h^2^=0.044, SE=0.048, p=0.18).

## Discussion

Here, we characterised DNA methylation outlier burden, which has been suggested as a promising new marker of biological age, in three large cohorts: The cross-sectional Generation Scotland cohort of individuals aged 18 to 92 years, and the Lothian Birth Cohorts of 1921 and 1936, which are longitudinal studies of older adults aged, on average, 79 and 69 years at baseline, respectively. We analysed methylation levels at those sites present on the Illumina Infinium HumanMethylation450k array, to permit comparison with findings by Wang and colleagues (13) in the Swedish Adoption/Twin Study of Aging (SATSA), a longitudinal cohort of older individuals in their 70s.

### DNA methylation outlier burden

There was high variability in DNA methylation outlier number between individuals. DNA methylation outlier burden was highly positively skewed and consequently, a log10-transformed version of outlier burden used in all analyses. This is consistent with previous findings (13,14).

When looking at outlier burden per CpG site rather than per individual, Wang and colleagues (13) reported that at 64% of CpG sites, at least one out of 385 individuals in SATSA had an outlier. Here, we found a similar number of CpG sites (63%) that were an outlier in at least one out of the 340 individuals in the LBC1921. However, our findings demonstrate that with increasing sample size, almost all sites will be an outlier at least once (83% of CpG sites in 898 individuals in the LBC1936 and >96% in the 7,010 individuals in Generation Scotland). This may suggest that the accumulation of DNA methylation outliers with age is a stochastic process, affecting most CpG sites randomly.

### DNA methylation outlier burden and age

DNA methylation outlier burden was found to be positively associated with age in Generation Scotland and in the Lothian Birth Cohorts. The association between age and log10(outlier burden) was significant in Generation Scotland independent of the level of adjustment and in the LBC1921 after adjustments for cell counts. A similar association was seen in the LBC1936 although it was not significant. Effect sizes were comparable to those reported by Wang and colleagues (β=8.3×10^−3^, p=1.2×10^−13^, (13)).

DNA methylation outlier burden measures were strongly associated with cell proportions and other technical factors, again, confirming previous findings (13,14). Moreover, in this study, batch effects were confounded by wave of data collection in the LBC1921 and LBC1936. Outliers were thus defined as values +/-3 interquartile ranges from the upper/lower quartile in each wave rather than at baseline, in order to reduce this confounding effect and obtain more stable definitions.

There was no difference in DNA methylation outlier burden between men and women in any of the three cohorts. This is consistent with findings reported by Curtis and colleagues (14) but not with those by Wang and colleagues (13) who found women to have a slightly higher outlier burden than men. To minimise confounding by sex, Wang and colleagues consequently defined outliers according to methylation level variability in women and men separately.

### DNA methylation outlier burden and epigenetic drift

We hypothesised that rather than merely being a consequence of an increase in outlier burden with age, the age-related increase in DNA methylation variability (epigenetic drift) may in fact contribute to the identification of larger numbers of outliers. An older individual’s methylation level might be classified as an outlier when compared to methylation levels in the entire sample, but it may not when compared to that of other older individuals (amongst whom variability is higher) (Supplementary Figure 5, Additional File 2).

Here, we found that controlling for epigenetic drift, by defining outliers based on variability in age deciles, only slightly attenuated the association between DNA methylation outlier burden and age in Generation Scotland.

Of course, defining outliers in even narrower age ranges could have further attenuated the association between outlier burden and age. However, there also was an association between outlier burden and age in the Lothian Birth Cohorts, despite outliers being defined within waves of data collection with very narrow age ranges. Taken together, these findings suggest that the age-related increase in extreme methylation levels (that is, epigenetic outliers), and the age-related increase in methylation level variability (epigenetic drift) appear to be, at least in part, two separate phenomena.

This is also consistent with findings by Wang and colleagues (13) who demonstrated that CpG sites where DNA methylation outliers occur frequently do not tend to be the same CpG sites where methylation level variability increases with age. Of 1,185 CpG sites with outliers in more than 50 individuals, only two had previously been identified as being variably methylated.

### DNA methylation outlier burden, health and survival

In contrast to previous studies, we did not find DNA methylation outlier burden to be associated with self-reported or family history of cancer (7,13). Furthermore, it was not associated with a comprehensive list of other self-reported disease outcomes. While other work has linked extreme DNA methylation patterns to congenital abnormalities (9) and low birth weight (15), the findings of the present study do not support strong links between DNA methylation outlier burden and age-related health outcomes.

In addition, we found mixed evidence regarding the association of DNA methylation outlier burden and health-related variables. DNA methylation outlier burden has previously been linked to smoking (16) but not to BMI (12,16). In Generation Scotland, higher DNA methylation outlier burden was found to be significantly positively associated with smoking pack years and significantly negatively with BMI. There were no significant associations between these variables in the LBC1921 and LBC1936.

Finally, there was a suggestive association between higher DNA methylation outlier burden and a higher risk of mortality.

### Heritability of DNA methylation outlier burden

Consistent with findings in twins by Wang and colleagues (13), there was no evidence that outlier burden is heritable.

### Strengths and limitations

Here, we provide a comprehensive characterisation of DNA methylation outlier burden in three large independent cohorts. The sample size of the Generation Scotland cohort is an order of magnitude greater than previous studies (12–14). In addition, this is the first study to systematically relate DNA methylation outlier burden to a range of diseases and to survival.

There are limitations to this study. First, the distribution of log10(outlier burden) in Generation Scotland was still slightly positively skewed after log10 transformation and its association with age was thus non-linear. Disease status, including cancer information, in the Generation Scotland study was measured by self-report which can be unreliable. In addition, this data was cross-sectional and for some diseases prevalence rates were low (e.g. Alzheimer’s disease and Parkinson’s disease; Supplementary Tables 5 and 6, Additional File 1) which may have limited the present study’s ability to detect significant associations. Finally, we did not run analyses separately on outliers based on their direction, that is, ‘high’ outliers with abnormally high methylation levels and ‘low’ outliers with lower than average methylation levels (13). We also did not investigate the genomic regions or functional pathways in which outliers may cluster (some work on this reported in 13,14,16).

## Conclusions

The findings of our study are consistent with previous work, demonstrating an increase in the number of DNA methylation outliers in individuals as they age. This suggests that DNA methylation outliers are unlikely to be just technical artefacts. Furthermore, our findings show that the accumulation of DNA methylation outliers with age is unlikely to be an artefact of epigenetic drift and that it does indeed appear to be driven by stochastic processes. We also demonstrate that the measurement of DNA methylation outliers is heavily associated with cell counts and technical factors. We do not find DNA methylation outlier burden to be robustly associated with age-related diseases and future studies in larger samples are needed to confirm whether it can predict survival. Based on the findings reported here, DNA methylation outlier burden may be useful as a marker of biological age and predict survival, but more work will be needed to establish whether DNA methylation outliers can offer insights into the associations between ageing, health and lifestyle.

## Methods

Here, we used data from the cross-sectional Generation Scotland cohort, as well as from the longitudinal Lothian Birth Cohorts of 1921 and 1936.

### The Generation Scotland cohort

Generation Scotland is a family-based cohort consisting of individuals aged 18 to 98 years, living across Scotland. Participants were initially recruited from individuals registered at GP surgeries and then asked to invite first degree relatives to join the study, resulting in a final sample size of 23,960 individuals. For these participants, genetic as well as clinical, lifestyle and sociodemographic information are available. Further details on the cohort can be found elsewhere (17,18).

The GS:SFHS has been ethically approved and granted research tissue bank status by the NHS Tayside Committee on Medical Research Ethics (REC reference numbers: 05/S1401/89 and 10/S1402/20, respectively). All study participants provided informed written consent.

### The Lothian Birth Cohorts of 1921 and 1936

The Lothian Birth Cohorts are two longitudinal cohort studies of ageing in individuals born in 1921 (LBC1921, n=550) or in 1936 (LBC1936, n=1,091). At age 11, as part of the Scottish Mental Health Surveys of 1932 and 1947, respectively, most of these individuals completed the Moray House Test of general intelligence. Decades later, those living in Edinburgh and the surrounding Lothian region were re-contacted and invited to participate in the Lothian Birth Cohort studies. Recruitment and baseline testing (wave 1) took place between 1999 and 2001 (mean age ∼ 79), and between 2004 and 2007 (mean age ∼ 70) for the LBC1921 and LBC1936, respectively. Since then, detailed physical, cognitive, psychosocial, and lifestyle information was collected roughly every 3 years in 4 subsequent waves of testing. In addition, genetic and longitudinal epigenetic profiling is available in both the LBC1921 and the LBC1936. More detail on recruitment and testing can be found elsewhere (19,20).

Ethical approval for the first wave of Lothian Birth Cohort studies was obtained from the Multi-Centre Research Ethics Committee for Scotland (MREC/01/0/56) and the Lothian Research Ethics committee (LREC/1998/4/183; LREC/2003/2/29). All participants provided written informed consent.

### DNA methylation data

Genome-wide DNA methylation in Generation Scotland was measured from blood samples collected between 2006 and 2011 (at the time of baseline appointment) using the Illumina Infinium HumanMethylationEPIC BeadChip at >850,000 CpG sites. The methylation profiling was carried out in two sets, here referred to as set 1 and set 2. Set 1 consisted of 5,200 individuals (2,586 of whom were genetically unrelated to each other at a relatedness threshold of <0.025); set 2 consisted of a further 4,683 unrelated individuals – both to others in set 2 and all of those in set 1. Methylation profiling in set 1 and set 2 was carried out in 31 batches each. Full details of DNA methylation quality control steps can be found in Additional File 2 (Supplementary Note 1).

DNA methylation in the Lothian Birth Cohorts was measured repeatedly using the Illumina HumanMethylation450K array at > 450,000 CpG sites. Methylation data are available in three waves in LBC1921 (wave 1, 3 and 4) and in four waves in LBC1936 (wave 1, 2, 3 and 4). Samples in LBC1936 were processed in three separate batches (Supplementary Table 8, Additional File 2) on 13 dates, using 41 plates and 309 microarrays in total. All samples in LBC1921 were processed in one batch on 7 dates, using 11 plates and 76 microarrays. Sample collection and quality control steps have been described in greater detail elsewhere (21,22). For a brief description, see Supplementary Note 1, Additional File 2.

Following quality control, methylation β-values were calculated using the m2beta function in *lumi* (23). Probes targeting polymorphic sites in the European population, ch-probes and probes predicted to hybridise to multiple genomic regions (identified for the EPIC array by McCartney et al., 2016) were removed. In addition, probes on the X and Y chromosomes and probes with known meQTLs were excluded. Note that the meQTL probes (both *cis* and *trans*) were identified using the Illumina Infinium HumanMethylation450 array (25) and that no comprehensive list is currently available for the EPIC array.

Following post-QC filtering, the datasets comprised the following sites and samples: 356,631 sites in 436 samples in the LBC1921 and in 906 samples in the LBC1936; 678,519 sites in 2,586 unrelated individuals in Generation Scotland set 1; and 724,207 sites in 4,450 individuals in Generation Scotland set 2. Here, we limit our analyses to the 334,352 and 349,027 sites measured in Generation Scotland set 1 and set 2, respectively, which are also present on the HumanMethylation450 array. This will make results from Generation Scotland more comparable to those obtained in the Lothian Birth Cohorts.

### Covariates

In addition to technical factors (batch and set in Generation Scotland; date, plate and array in the Lothian Birth Cohorts), a number of phenotypes were included as covariates in the analyses. These included white blood cell proportions of granulocytes, natural killer (NK) cells, B-, CD4^+^T- and CD8^+^T-cells which had been estimated from DNA methylation using the Houseman method (26) as implemented in the *estimateCellCounts* function in *minfi* (27). Furthermore, a binary ever-never smoking variable, a log10-transformed continuous smoking pack years variable, a binary self-report cancer diagnosis (Yes/ No) variable and a log10-transformed continuous measure of body mass index (BMI) were included as covariates. Variables were log10-transformed in order to minimise positive skew. Full details can be found in Supplementary Notes 1 and 2, Additional File 2.

### Health and survival data

In Generation Scotland, DNA methylation outlier burden was related to more than a dozen binary measures of self-reported disease and self-reported family history of disease. In the Lothian Birth Cohorts, DNA methylation outlier burden was related to survival. Mortality data was provided by the General Register Office for Scotland (National Records Scotland) through data linkage to the National Health Service Central Register. The data used in the analyses reported here are correct as of January 2018 (LBC1921) and April 2018 (LBC1936). At time of last censor (approximately 18 years from baseline for the LBC1921 and approximately 14 years from baseline for the LBC1936), 37 and 680 participants of the LBC1921 and LBC1936 were alive, respectively.

### DNA methylation outlier burden

In Generation Scotland, DNA methylation outliers were defined as per Gentilini and colleagues (12), that is, as sites with methylation levels greater than three times the interquartile range (IQR) from the upper or lower quartile. Outliers were calculated within each set separately. Data from Generation Scotland set 1 and set 2 were then combined for all following analyses. In the longitudinal Lothian Birth Cohort studies, DNA methylation outliers were calculated in each wave of data collection based on the IQR calculated in that wave (rather than the IQR in wave 1, as done by Wang and colleagues (13)). The reason for this is that there are strong technical effects on DNA methylation measures, and that in the Lothian Birth Cohorts, technical factors are confounded by wave of sample collection. For instance, set is confounded by wave of sample collection in the LBC1936 (Supplementary Table 8, Additional File 1). In addition, factors such as plate and date are confounded by wave even within processing sets. By defining outliers within each wave, we hoped to minimise these batch effects, resulting in more stable outlier definitions. Finally, outlier burden per individual was calculated as the total number of outlier sites for each individual. In addition, outliers per CpG site were calculated as the total number of individuals with an outlier per site.

Following this, samples with extreme values for any of the estimated cell proportions (> 5 standard deviations from the mean) were removed (5 and 13 samples in Generation Scotland set 1 and set 2, and 7 and 32 samples in LBC1921 and LBC1936, respectively). In addition, a further eight individuals from Generation Scotland set 1 were excluded from the analyses because they had responded with ‘Yes’ to every self-reported disease phenotype included on the questionnaire.

A combined dataset of 7,010 individuals remained for analysis in Generation Scotland (2,573 in set 1 and 4,437 in set 2). The final Lothian Birth Cohort samples consisted of 430 individuals (685 measurements) in the LBC1921 and 898 individuals (2801 measurements) in the LBC1936.

### Analyses

All analyses were performed in R (28). The *lme4* (29), *lmerTest* (30) and *survival* (31) packages were used to construct linear mixed models and Cox regression models. The *ggplot2* (32) and *survminer* (33) packages were used to create graphs. For a flowchart displaying the analysis steps, see Figure 4.

**Figure 4.**
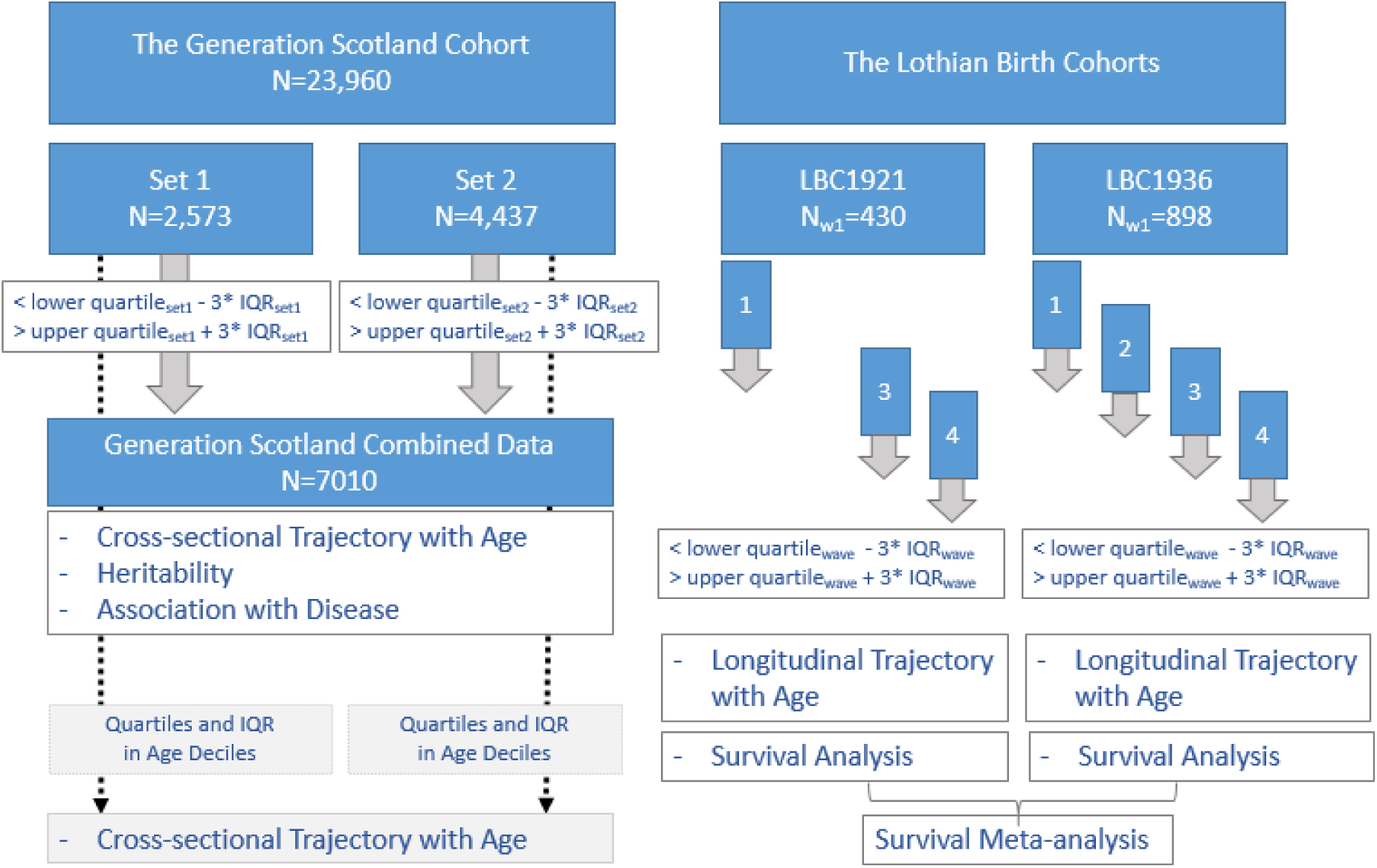
Flowchart of analyses in the Generation Scotland and Lothian Birth Cohorts. Methylation profiling in Generation Scotland was carried out in two separate sets. DNA methylation outliers were calculated within each set. Analyses were then carried out on a combined dataset. In the Lothian Birth Cohorts of 1921 and 1936, DNA methylation outliers were calculated within each wave of data collection. Analyses were carried out in the LBC1921 and the LBC1936 separately. Results from the survival analyses in both cohorts were meta-analysed.

#### DNA methylation outlier burden and age

Cross-sectional associations between a log10-transformed version of DNA methylation outlier burden and age in Generation Scotland were obtained by fitting three linear models. The basic model included sex as a covariate. The second model included additional adjustments for estimated white blood cell proportions (granulocytes, natural killer (NK) cells, B-, CD4^+^T- and CD8^+^T-cells) and technical variables (batch and set). The third (fully adjusted) model also controlled for smoking (ever/never), log10(pack years + 1), cancer diagnosis (Yes/No), and log10(BMI). The first model was a linear regression, whereas models 2 and 3 were linear mixed effects models with a random intercept term fitted for batch.

Longitudinal analyses of outlier burden in the Lothian Birth Cohorts were run by fitting the following three linear mixed models with random intercepts for ID and batch variables. The most basic model included sex as a fixed effect. The adjusted model included sex, estimated white blood cell proportions (granulocytes, natural killer (NK) cells, B-, CD4^+^T- and CD8^+^T-cells) and technical factors (date, plate and array) with final adjustments made for smoking (ever/never), log10(pack years + 1), cancer diagnosis (Yes/No), and log10(BMI).

#### DNA methylation outlier burden and epigenetic drift

Variability in methylation levels is known to increase over time (epigenetic drift) (5,6). In order to investigate whether this affects the association between outlier burden and age, we re-calculated outliers in Generation Scotland in an alternative fashion. We defined outliers as extreme methylation levels with respect to variability between individuals of the same age group rather than variability overall (Supplementary Figure 5, Additional File 2). We defined outliers as methylation levels greater than three times the IQR above or below the upper and lower quartile based on the variability within age deciles.

#### DNA methylation outlier burden, health and survival

Logistic regression models with basic adjustments for age and sex, followed by full adjustment for cell count proportions, smoking status and log(pack years), cancer, and log(BMI) were fit to investigate the association between DNA methylation outlier burden and binary measures of self-reported disease in the large Generation Scotland dataset. As prevalence rates for some diseases were low, analyses were repeated using self-reports of family history (in mother or father) of disease. We corrected for multiple testing using Bonferroni correction (p<0.05/13=3.9×10^−3^ for self-reported disease and p<0.05/16=3.1×10^−3^ for self-reported family history of disease).

Multivariate Cox proportional hazards models with two levels of adjustments (as per the logistic models described above) were fit to study the association between survival and outlier burden at baseline in the Lothian Birth Cohorts of 1921 and 1936.

A meta-analysis of hazard ratios obtained in the LBC1921 and the LBC1936 was run using a fixed effect model as implemented in the rma.uni function in the *metafor* package (34).

#### Heritability of DNA methylation outlier burden

Genotype data at >700,000 SNPs in Generation Scotland was generated using the Illumina OMNIExpress chip. Briefly, SNPs with more than 2% missingness and a Hardy-Weinberg Equilibrium test p<10^−6^ were excluded during quality control. This has been described in greater detail elsewhere (35). The genetic contribution by all genome-wide SNPs to phenotypic variance in log10(outlier burden) was estimated using Genome-wide Complex Trait Analysis (GCTA), GCTA-GREML (36).

## Data Availability

According to the terms of consent for Generation Scotland participants, access to data must be reviewed by the Generation Scotland Access Committee. Applications should be made to.
Lothian Birth Cohort data are available on request from the Lothian Birth Cohort Study, Centre for Cognitive Ageing and Cognitive Epidemiology, University of Edinburgh. Lothian Birth Cohort data are not publicly available due to them containing information that could compromise participant consent and confidentiality.

## Declarations

### Ethics approval and consent to participate

The GS:SFHS has been ethically approved and granted research tissue bank status by the NHS Tayside Committee on Medical Research Ethics (REC reference numbers: 05/S1401/89 and 10/S1402/20, respectively). Ethical approval for the first wave of Lothian Birth Cohort studies was obtained from the Multi-Centre Research Ethics Committee for Scotland (MREC/01/0/56) and the Lothian Research Ethics committee (LREC/1998/4/183; LREC/2003/2/29). All participants provided written informed consent.

### Consent for publication

Not applicable.

### Availability of data and materials

According to the terms of consent for Generation Scotland participants, access to data must be reviewed by the Generation Scotland Access Committee. Applications should be made to.

Lothian Birth Cohort data are available on request from the Lothian Birth Cohort Study, Centre for Cognitive Ageing and Cognitive Epidemiology, University of Edinburgh. Lothian Birth Cohort data are not publicly available due to them containing information that could compromise participant consent and confidentiality.

### Competing interests

AMM has received research support from Eli Lilly, Janssen and the Sackler Trust. AMM has also received speaker fees from Janssen and Illumina. The other authors declare that they have no competing interests.

### Funding

GS:SFHS is funded by the Scottish Government Health Department. In addition, Generation Scotland was developed with the support of Scottish Enterprise and receives funding and support from the Scottish Funding Council and the Chief Scientist Office of the Scottish Government. STRADL is funded by the Wellcome Trust (Reference 104036/Z/14/Z).

The LBC1921 was supported by the UK’s Biotechnology and Biological Sciences Research Council (BBSRC), a Royal Society–Wolfson Research Merit Award to IJD, and the Chief Scientist Office (CSO) of the Scottish Government’s Health Directorates. The LBC1936 is supported by Age UK (Disconnected Mind program) and the Medical Research Council (MR/M01311/1). Methylation typing was supported by Centre for Cognitive Ageing and Cognitive Epidemiology (Pilot Fund award), Age UK, The Wellcome Trust Institutional Strategic Support Fund, The University of Edinburgh, and The University of Queensland.

This work was in part conducted in the Centre for Cognitive Ageing and Cognitive Epidemiology, which is supported by the Medical Research Council and Biotechnology and Biological Sciences Research Council (MR/K026992/1).

AS is supported by a Medical Research Council PhD Studentship in Precision Medicine with funding by the Medical Research Council Doctoral Training Programme and the University of Edinburgh College of Medicine and Veterinary Medicine. REM and DLM are supported by Alzheimer’s Research UK major project grant ARUK-PG2017B-10. SH is supported by Loo and Hans Osterman Foundation, Foundation for geriatric diseases, Magnus Bergwall foundation, Foundation Gamla Tjänarinnor, Erik Rönnberg award for scientific studies on ageing and age-related diseases, King Gustaf Vs and Queen Victorias Freemanson Foundation for aging studies, Karolinska Institutet (Strategic Research Area in Epidemiology, KID), the Swedish Research Council (2015-03255) and the Swedish Council for Working Life and Social Research (FORTE) (2013-2292). RFH and AJS are supported by funding from the Wellcome Trust 4-year PhD in Translational Neuroscience–training the next generation of basic neuroscientists to embrace clinical research [108890/Z/15/Z].

### Authors’ contributions

Conception and design: AS and REM. Data analysis: AS and DLM. Interpretation: AS, YW, SH, IJD and REM. Drafting the article: AS and REM. Revision of the article: All other authors.

## Acknowledgements

The authors thank all individuals and project team members who have contributed to both GS:SFHS and to the ‘STRADL: Stratifying Resilience and Depression Longitudinally’ follow-up study. The authors also thank all LBC1921 and LBC1936 study participants and research team members who have contributed, and continue to contribute, to ongoing studies.

